# Adverse outcomes associated with Long-Term Opioid Therapy Discontinuation in People with Chronic Non-Cancer Pain in UK Primary Care: A propensity Score Matched Cohort Study

**DOI:** 10.1101/2025.03.21.25324373

**Authors:** Qian Cai, Yun-Ting (Joyce) Huang, Christos Grigoroglou, Charlotte Morris, Thomas Allen, Evangelos Kontopantelis

## Abstract

**Objective:** To examine adverse outcomes associated with long-term opioid therapy (L-TOT) discontinuation among people with chronic non-cancer pain (CNCP).

**Design and setting:** Population-based matched cohort study using UK Clinical Practice Research Datalink Aurum data.

**Population:** L-TOT discontinuers were defined as individuals with opioid-free for ≥180 days following an episode of L-TOT (≥3 opioid prescriptions within 90 days, or total ≥90 supply days within the first year, excluding the initial 30 days) between 01/01/2000-31/12/2020.

**Main outcome measures:** Propensity-score matched (1:5) cohorts of L-TOT discontinuers and L-TOT users were created to assess the association between L-TOT discontinuation and opioid-related death, hospitalisations due to bone fractures, their composite and all-cause mortality, using adjusted Cox regression models. If proportional hazards assumptions were violated, time-dependent Cox models were applied.

**Results:** A total of 29,589 L-TOT discontinuers (female 60.04%, mean age 56.45±18.42 years) were identified and matched with L-TOT users. The median follow-up durations were 6.87 years (interquartile range: 3.32 to 11.51) for opioid-related death in L-TOT discontinuers, compared to 5.74 years (2.51 to 10.18) in L-TOT users. Similar follow-up durations were observed for hospitalisations due to bone fractures, the composite outcome, and all-cause mortality. After adjusting for covariates used in the propensity score models, L-TOT discontinuation was associated with a 37% reduced risk in opioid-related death (adjusted hazards ratio: 0.63, 95% confidence interval: 0.42 to 0.94), a 5% reduction in the risk of hospitalisations due to bone fractures (0.95, 0.91 to 0.99), a 7% reduced risk of the composite outcome (0.93, 0.87 to 0.97), and a 22% lower risk of all-cause mortality (0.78, 0.76 to 0.80).

**Conclusions:** L-TOT discontinuation was associated with reduced risks of opioid-related death, hospitalisations due to bone fractures and all-cause mortality, suggesting important safety benefits of this practice. Further research should assess broader health outcomes beyond those examined in this study.

**What is already known on this topic:**

- Despite safety and effectiveness concerns, a significant proportion of people (12.2%) in UK primary care rely on L-TOT to manage their CNCP.
- UK clinical guidelines recommend reducing or stopping L-TOT when potential harms outweigh the benefits.
- Previous studies, primarily from the US, have shown mixed outcomes of L-TOT discontinuation, with some reporting positive effects such as reduced pain severity, while others highlighting significant risks including increased risks of suicide, overdose, and mental health crises.

**What this study adds:**

- Our study found that in UK primary care, patients with CNCP who discontinued L-TOT had a 37% lower risk of opioid-related death, a 5% reduced risk of hospitalisations due to bone fractures, and a 22% lower risk of all-cause mortality.
- Our findings support research into transitioning from L-TOT to alternative non-opioid pain management strategies, as no significant risks of adverse outcomes were observed with discontinuation.

## Introduction

Although the use of long-term opioid therapy (L-TOT) for chronic non-cancer pain (CNCP) decreased from 2009 to 2019, a significant proportion of patients (12.2%) in UK primary care continue to rely on L-TOT for their pain management. ^1, 2^ Prolonged opioid use is associated with significant risks, including increased emergency department visits, ^3^ bone fractures, ^4, 5^ and opioid-related death. ^6, 7^ Recognising these dangers, UK clinical guidelines recommend reducing or stopping L-TOT when potential harms outweigh the benefits. ^8, 9^ To support safe L-TOT discontinuation, a clear understanding of the outcomes associated with L-TOT discontinuation is needed. However, this has been poorly investigated in UK primary care to date.

Current evidence on the outcomes of L-TOT discontinuation primarily comes from United States (US)-based studies, with mixed findings. Some studies reported positive effects of stopping or reducing L-TOT, such as reduced pain severity ^10^ and improvements in physical function and quality of life. ^11^ Others highlighted significant risks associated with discontinuation, including increased rates of suicide, ^12, 13^ overdose, ^13, 14^ mental health crises, ^15^ and death. ^16^ These adverse outcomes are particularly pronounced among patients prescribed high daily opioid doses (≥120 morphine milligram equivalents [MME]), those with a history of mental health conditions, or those concurrently using benzodiazepines. This conflicting evidence has led to uncertainty about whether discontinuing L-TOT is ultimately beneficial or harmful, especially given the complex comorbidities and high vulnerabilities often seen in people with CNCP.

Despite these insights, evidence on the outcomes of L-TOT discontinuation in the UK remains limited. The risks and benefits of discontinuation may be influenced by differences in healthcare systems, prescribing practices, patient management approaches, and opioid utilisation patterns, which vary significantly from those in the US. While previous US-based studies have examined several adverse outcomes associated with L-TOT discontinuation, other critical outcomes, such as the risk of bone fractures, have received little attention. L-TOT is known to increase the risk of falls and reduced bone density, ^17^ and these effects may either attenuate or persist after discontinuation; however, the extent to which discontinuation modifies fracture risk remains unclear. Additionally, previous studies have not fully assessed the broader impact of discontinuation on opioid-related mortality and all-cause mortality within the UK context. To address these gaps, this study aimed to assess the association between L-TOT discontinuation and key adverse outcomes, including opioid-related death, hospitalisations due to bone fractures, their composite, and all-cause mortality among people with CNCP in UK primary care. By providing robust evidence on these risks, we seek to inform clinical decision-making and guide policies that enhance patient safety while mitigating potential harms.

## Methods

### Study design and population

The study design and population was described in our previous work. ^1^ Briefly, in this population-based matched cohort study, we used data from the Clinical Practice Research Datalink (CPRD) Aurum, a database of anonymised UK primary care electronic health records, ^18^ covering 20% of the national population (approximately 13 million active registrants). We included people aged ≥18 years on the index date (the first opioid prescription following a 12-month period of opioid-free) with a diagnosis of CNCP (code list is provided in Supplementary Table S1) between 1st January 2000 and 31st December 2020. Among them, we identified L-TOT users, defined as individuals receiving ≥3 opioid prescriptions within a 90-day period, or a sum of opioid supply days lasting ≥90 days, within the first year of follow-up (excluding the first 30 days) ^19^, and L-TOT discontinuers, defined as those had no opioid records for ≥180 days following an episode of L-TOT. CNCP could occur up to 6 months after or any time before the index date. ^20^ Patients with a cancer diagnosis, except for non-melanoma skin cancer, in the previous ten years were excluded due to different drug utilisation patterns and different guidelines for opioid prescribing. To be eligible, patients were required to have been registered with the CPRD practice for at least 12 months and have available linked data for the Index of Multiple Deprivation (IMD) 2019, ^21^ a measure of socioeconomic status in England; the Hospital Episode Statistics Admitted Patient Care (HES APC) database, which contains records of hospital admissions; and the Office for National Statistics (ONS) mortality data, which records death registrations.

### Follow-up period

For L-TOT users, follow-up began from either the end date of the last opioid prescription within the qualified 90-day period; or the end date of the last opioid prescription contributing to the cumulative supply of ≥90 days, until the earliest occurrence of an outcome of interest (specified in the Outcomes section), transfer out of the general practice, the last data collection date of the general practice, a cancer diagnosis, or the end of the study (31 December 2020). For L-TOT discontinuers, follow-up began on the date of the next recorded opioid prescription following a gap of ≥180 days without opioid use after an episode of L-TOT, until the same endpoints as described for L-TOT users. The study cohort identification process is shown in Supplementary Figure S1.

### Opioid use

All non-parental opioid analgesics (code list is provided in Supplementary Table S2) were included, except for methadone and sublingual buprenorphine, as their primary indication is for opioid addiction rather than pain management. Opioid drug exposure data were processed using the logic of a previously published drug preparation algorithm. ^19, 22^ Details of the relevant decisions were provided in Supplementary Figure S2. Opioid doses were converted to oral morphine milligram equivalents (MME) using standardised conversion factors. ^23^

### Outcomes

We examined opioid-related death, hospitalisations due to bone fractures, their composite (i.e., either having opioid-related death or hospitalisations due to bone fractures, or both) and all-cause mortality. Hospitalisations due to bone fractures was identified using ICD-10 (international classification of diseases, 10^th^ Version) codes (Supplementary Table S3) from the linked HES APC database, while opioid-related death and all-cause mortality data were obtained from the ONS death registration records, also using relevant ICD-10 codes (Supplementary Table S4). We used the first record of these outcomes during the follow-up period, which means that patients had events prior to the start of follow-up were excluded from the analysis for this specific outcome. This approach was taken to avoid potential confounding, particularly for hospitalisations due to bone fractures, as a history of bone fracture is associated with a higher risk of recurrence, ^24, 25^ which may lead to hospitalisations. However, these patients remained eligible for analyses of other outcomes.

### Covariates

Baseline patient characteristics (i.e., age, sex) were collected on the index date. Ethnicity data was obtained through the linkage with HES APC data. ^26^ Geographical regions included North East, North West, Yorkshire & the Humber, East Midlands, West Midlands, East of England, South West, South Central, South East Coast and London. Socioeconomic status was measured using the IMD 2019 ^21^ and grouped into quintiles (Q1-least deprived, Q5-most deprived). Lifestyle factors included smoking and alcohol drinking status and were categorised in four groups: never, current, former, unknown.

Comorbidities were measured using Charlson comorbidity index (CCI) score and categorised into three grades: low (0-2); medium (3-4); and high (≥5). ^27^ The presence of other conditions, including substance use disorders, alcohol dependence, anxiety, depression, schizophrenia, osteoarthritis, rheumatoid arthritis, and epilepsy were identified within five years prior to the index date. We also identified concurrent medication use including benzodiazepines, gabapentinoids, antidepressants, Z-drugs, muscle relaxants or non-steroidal anti-inflammatory drugs (NSAIDs) within five years prior to the index date.

### Statistical analysis

#### Crude incidence rates

The crude incidence rate (95% confidence interval [CI]) of each outcome was calculated before matching, by dividing the total number of new events by total person-years at risk, separately for L-TOT discontinuers and L-TOT users.

#### Propensity-score matched cohort

L-TOT discontinuers were matched with L-TOT users in a 1:5 ratio using a propensity score matching (PSM) approach with replacement. Propensity scores were estimated using multivariable logistic regression, with a calliper width of 0.02. L-TOT discontinuation was the dependent variable in the logistic regression model, with all covariates mentioned above included as independent variables. There were missing values for ethnicity, drinking status, IMD and smoking status, and they were categorised as ‘unknown’ and included in the propensity score (PS) models. Although we initially created a single matched cohort, the eligibility criteria differed for each outcome, particularly for hospitalisations due to bone fractures. Patients with a history of fracture-related hospitalisation prior to follow-up were excluded from that specific analysis but remained eligible for opioid-related death and all-cause mortality. Therefore, to account for these differences in cohort composition, we conducted separate PS models for each outcome. The effectiveness of PSM was evaluated by calculating standardised differences (SDs) for all covariates before and after matching, with SDs less than 10% considered good balance between the two groups. ^28^ We also plotted kernel density estimates to assess the balance in propensity scores of the two groups before and after matching (Supplementary Figure S3).

#### Adjusted Cox proportional hazards regressions

Cox proportional hazards regression model was used to estimate the risks of each outcome associated with L-TOT discontinuation, controlling for covariates that were used to create the PSM cohorts. Results were presented in adjusted hazard ratios (aHRs) with 95% CIs. Adjusted survival curves based on the Cox proportional hazards model were plotted to compare the predicted survival probabilities between L-TOT discontinuers and L-TOT users, accounting for covariates. The proportional hazards (PH) assumption was graphically assessed by plotting Schoenfeld residuals against time for each covariate and statistically evaluated using both global and covariate-specific tests. In models where the PH assumption was violated, a time-dependent Cox regression model was applied instead, incorporating covariates that drove the PH assumption violation as time-dependent (details in the Results section). Model performance was assessed using the Akaike Information Criterion (AIC) and Bayesian Information Criterion (BIC), with the lowest AIC/BIC values indicating the best fit. ^29^ To ensure the robustness of our findings, we further employed parametric survival models based on Weibull distribution as alternative analytical approaches when the PH assumptions were violated and compared their results with those obtained from time-dependent Cox regression models.

#### Sensitivity analyses

In sensitivity analyses, we matched L-TOT discontinuers and L-TOT users in a 1:1 ratio using the same PSM approach, applying the same covariates adjustment process described in the main analysis. We also conduced Fine-Gray subdistribution hazards regression models (i.e., the competing-risks regressions) using all-cause mortality as a competing event to examine the association between L-TOT discontinuation and the other three outcomes (i.e., opioid-related death, hospitalisations due to bone fractures, and their composite), while adjusting for the aforementioned covariates. The results were reported with subhazard ratios (sHRs) and 95%CI. All analyses were conducted using Stata/MP version 18.0.

### Patient and public involvement

We have engaged patient groups, including the PRIMER (Primary Care Research in Manchester Engagement Resource, https://sites.manchester.ac.uk/primer/) at the University of Manchester, to assist in developing our dissemination strategy.

## Results

### Patient characteristics

The number of L-TOT discontinuers and their matched L-TOT users included in the analysis varied slightly by outcome. Table 1 summarised the baseline characteristics of L-TOT discontinuers and L-TOT users for the analysis of opioid-related death before and after the 1:5 PSM. Before matching, a total of 29,589 L-TOT discontinuers (White 79.85%, female 60.04%, mean age 56.45±18.42 years) with CNCP were identified from 573,639 L-TOT users in the first year of follow-up between 1^st^ Jan 2000 and 31 Dec 2020 (Supplementary Figure S4). A 1:5 PSM was performed, resulting in a matched cohort of 148,109 individuals (White 82.55%, female 59.57%, mean age of 56.83±18.60 years) used in the analysis of the risk of opioid-related death. The distributions of propensity scores for the two groups (L-TOT discontinuers vs L-TOT users), as reflected by the density curves (Supplementary Figure S3), showed substantial overlap, indicating that the matching procedure successfully balanced the covariates. After matching, the two groups were more comparable in terms of age, daily MME, comorbidities (e.g., osteoarthritis, anxiety, alcohol dependence, epilepsy, substance use disorder), CCI, and concurrent NSAIDs use, with L-TOT discontinuers becoming more similar to L-TOT users descriptively. Baseline patient characteristics for the other three outcomes (i.e., hospitalisations due to bone fractures, the composite outcome and all-cause mortality) showed similar distributions to those reported for opioid-related death (Supplementary Tables S5-S7).

**Table 1.**
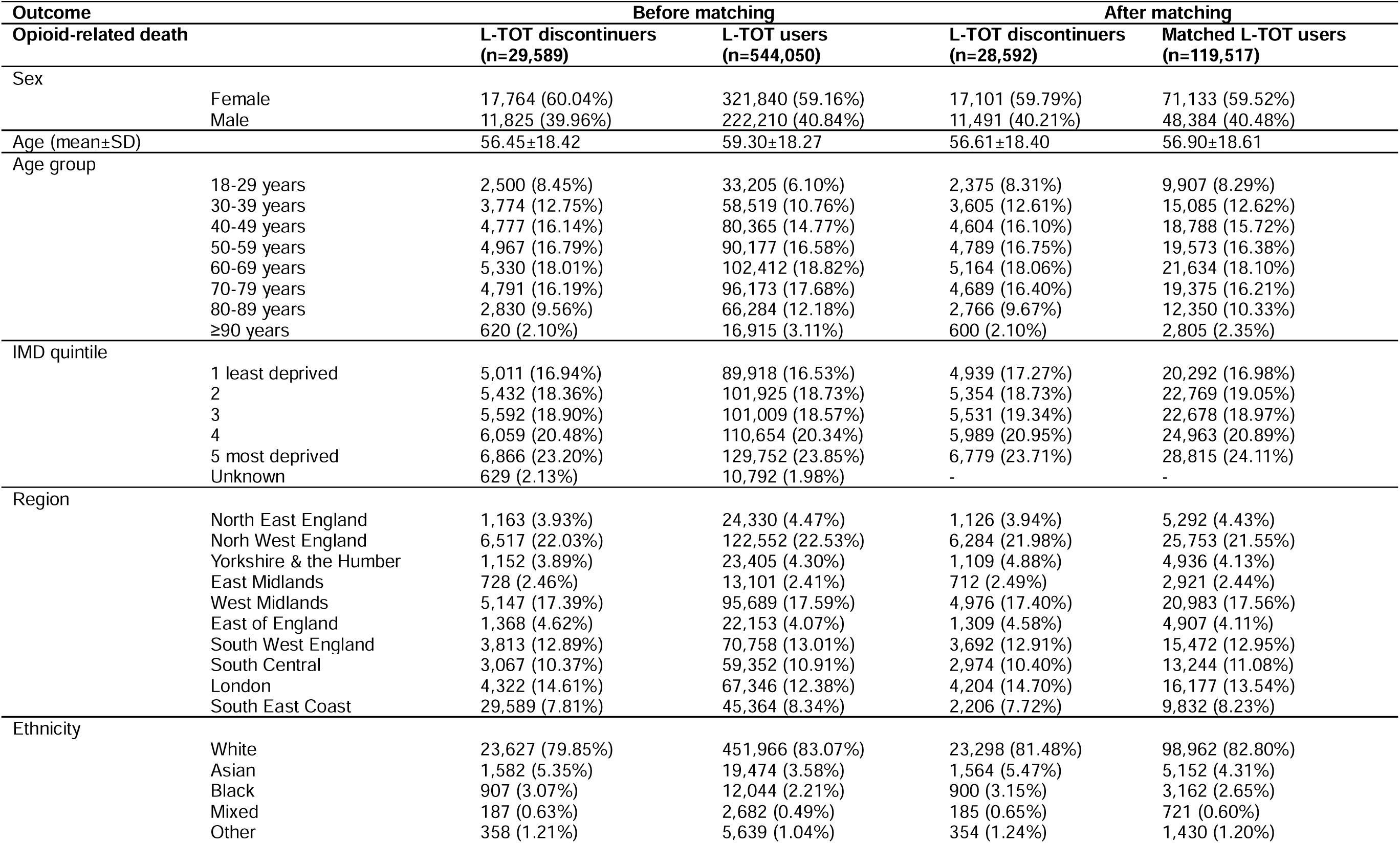
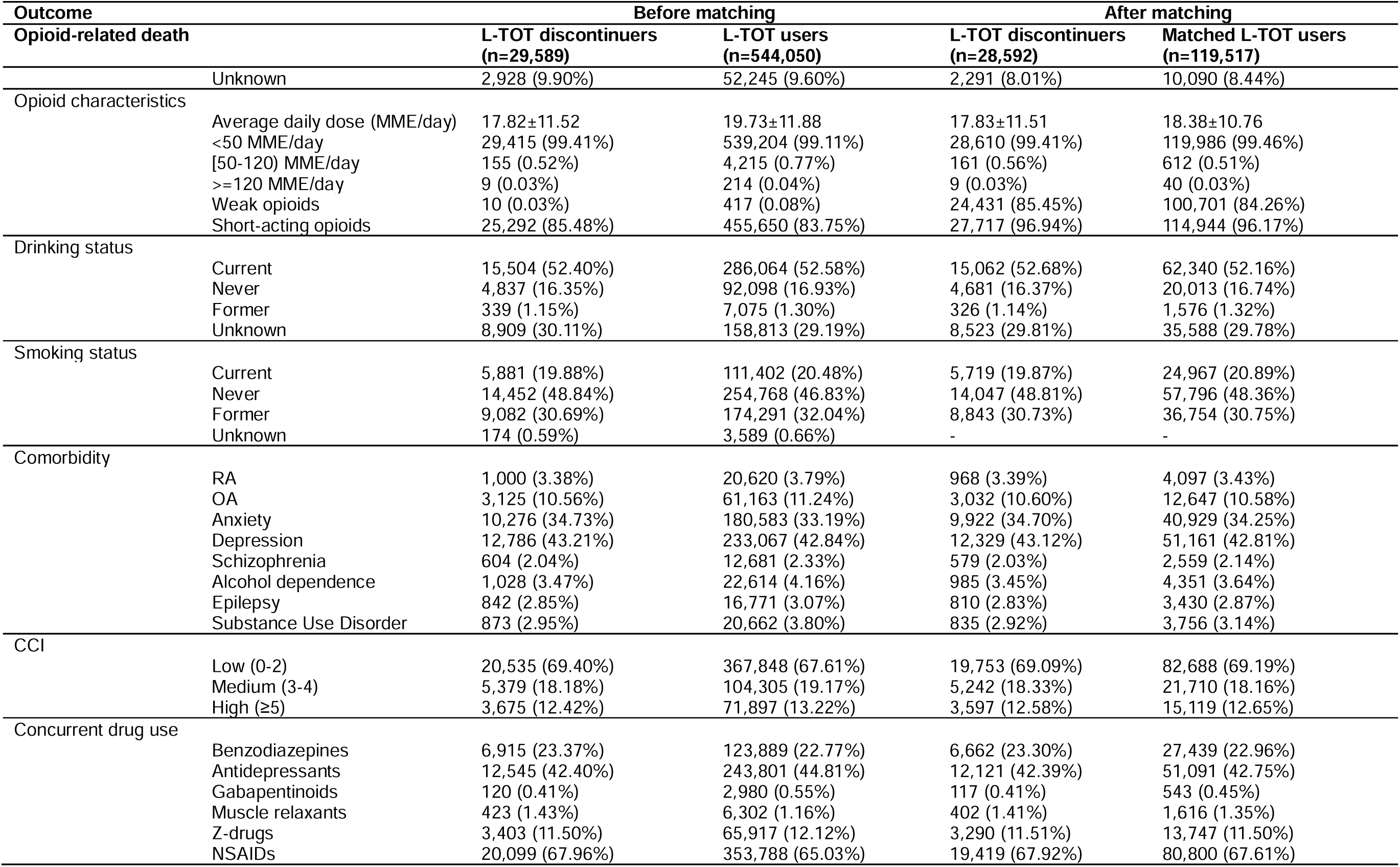
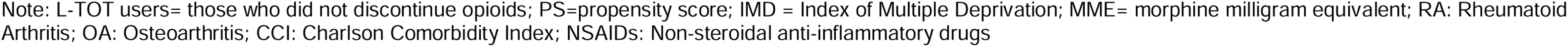
Baseline characteristics between L-TOT discontinuers and L-TOT users for the analysis of opioid-related death before and after 1:5 matching between 01/01/2000 - 31/12/2020.

| Outcome |  | Before matching |  | After matching |  |
| --- | --- | --- | --- | --- | --- |
| Opioid-related death |  | L-TOT discontinuers<br>(n=29,589) | L-TOT users<br>(n=544,050) | L-TOT discontinuers<br>(n=28,592) | Matched L-TOT users<br>(n=119,517) |
| Sex | Female | 17,764 (60.04%) | 321,840 (59.16%) | 17,101 (59.79%) | 71,133 (59.52%) |
|  | Male | 11,825 (39.96%) | 222,210 (40.84%) | 11,491 (40.21%) | 48,384 (40.48%) |
| Age (mean±SD) |  | 56.45±18.42 | 59.30±18.27 | 56.61±18.40 | 56.90±18.61 |
| Age group | 18-29 years | 2,500 (8.45%) | 33,205 (6.10%) | 2,375 (8.31%) | 9,907 (8.29%) |
|  | 30-39 years | 3,774 (12.75%) | 58,519 (10.76%) | 3,605 (12.61%) | 15,085 (12.62%) |
|  | 40-49 years | 4,777 (16.14%) | 80,365 (14.77%) | 4,604 (16.10%) | 18,788 (15.72%) |
|  | 50-59 years | 4,967 (16.79%) | 90,177 (16.58%) | 4,789 (16.75%) | 19,573 (16.38%) |
|  | 60-69 years | 5,330 (18.01%) | 102,412 (18.82%) | 5,164 (18.06%) | 21,634 (18.10%) |
|  | 70-79 years | 4,791 (16.19%) | 96,173 (17.68%) | 4,689 (16.40%) | 19,375 (16.21%) |
|  | 80-89 years | 2,830 (9.56%) | 66,284 (12.18%) | 2,766 (9.67%) | 12,350 (10.33%) |
|  | ≥90 years | 620 (2.10%) | 16,915 (3.11%) | 600 (2.10%) | 2,805 (2.35%) |
| IMD quintile | 1 least deprived | 5,011 (16.94%) | 89,918 (16.53%) | 4,939 (17.27%) | 20,292 (16.98%) |
|  | 2 | 5,432 (18.36%) | 101,925 (18.73%) | 5,354 (18.73%) | 22,769 (19.05%) |
|  | 3 | 5,592 (18.90%) | 101,009 (18.57%) | 5,531 (19.34%) | 22,678 (18.97%) |
|  | 4 | 6,059 (20.48%) | 110,654 (20.34%) | 5,989 (20.95%) | 24,963 (20.89%) |
|  | 5 most deprived | 6,866 (23.20%) | 129,752 (23.85%) | 6,779 (23.71%) | 28,815 (24.11%) |
|  | Unknown | 629 (2.13%) | 10,792 (1.98%) | - | - |
| Region | North East England | 1,163 (3.93%) | 24,330 (4.47%) | 1,126 (3.94%) | 5,292 (4.43%) |
|  | North West England | 6,517 (22.03%) | 122,552 (22.53%) | 6,284 (21.98%) | 25,753 (21.55%) |
|  | Yorkshire & the Humber | 1,152 (3.89%) | 23,405 (4.30%) | 1,109 (4.88%) | 4,936 (4.13%) |
|  | East Midlands | 728 (2.46%) | 13,101 (2.41%) | 712 (2.49%) | 2,921 (2.44%) |
|  | West Midlands | 5,147 (17.39%) | 95,689 (17.59%) | 4,976 (17.40%) | 20,983 (17.56%) |
|  | East of England | 1,368 (4.62%) | 22,153 (4.07%) | 1,309 (4.58%) | 4,907 (4.11%) |
|  | South West England | 3,813 (12.89%) | 70,758 (13.01%) | 3,692 (12.91%) | 15,472 (12.95%) |
|  | South Central | 3,067 (10.37%) | 59,352 (10.91%) | 2,974 (10.40%) | 13,244 (11.08%) |
|  | London | 4,322 (14.61%) | 67,346 (12.38%) | 4,204 (14.70%) | 16,177 (13.54%) |
|  | South East Coast | 29,589 (7.81%) | 45,364 (8.34%) | 2,206 (7.72%) | 9,832 (8.23%) |
| Ethnicity | White | 23,627 (79.85%) | 451,966 (83.07%) | 23,298 (81.48%) | 98,962 (82.80%) |
|  | Asian | 1,582 (5.35%) | 19,474 (3.58%) | 1,564 (5.47%) | 5,152 (4.31%) |
|  | Black | 907 (3.07%) | 12,044 (2.21%) | 900 (3.15%) | 3,162 (2.65%) |
|  | Mixed | 187 (0.63%) | 2,682 (0.49%) | 185 (0.65%) | 721 (0.60%) |
|  | Other | 358 (1.21%) | 5,639 (1.04%) | 354 (1.24%) | 1,430 (1.20%) |
| Opioid-related death |  | L-TOT discontinuers<br>(n=29,589) | L-TOT users<br>(n=544,050) | L-TOT discontinuers<br>(n=28,592) | Matched L-TOT users<br>(n=119,517) |
| Unknown |  | 2,928 (9.90%) | 52,245 (9.60%) | 2,291 (8.01%) | 10,090 (8.44%) |
| Opioid characteristics |  |  |  |  |  |
| Average daily dose (MME/day) |  | 17.82±11.52 | 19.73±11.88 | 17.83±11.51 | 18.38±10.76 |
| <50 MME/day |  | 29,415 (99.41%) | 539,204 (99.11%) | 28,610 (99.41%) | 119,986 (99.46%) |
| [50-120) MME/day |  | 155 (0.52%) | 4,215 (0.77%) | 161 (0.56%) | 612 (0.51%) |
| ≥120 MME/day |  | 9 (0.03%) | 214 (0.04%) | 9 (0.03%) | 40 (0.03%) |
| Weak opioids |  | 10 (0.03%) | 417 (0.08%) | 24,431 (85.45%) | 100,701 (84.26%) |
| Short-acting opioids |  | 25,292 (85.48%) | 455,650 (83.75%) | 27,717 (96.94%) | 114,944 (96.17%) |
| Drinking status |  |  |  |  |  |
| Current |  | 15,504 (52.40%) | 286,064 (52.58%) | 15,062 (52.68%) | 62,340 (52.16%) |
| Never |  | 4,837 (16.35%) | 92,098 (16.93%) | 4,681 (16.37%) | 20,013 (16.74%) |
| Former |  | 339 (1.15%) | 7,075 (1.30%) | 326 (1.14%) | 1,576 (1.32%) |
| Unknown |  | 8,909 (30.11%) | 158,813 (29.19%) | 8,523 (29.81%) | 35,588 (29.78%) |
| Smoking status |  |  |  |  |  |
| Current |  | 5,881 (19.88%) | 111,402 (20.48%) | 5,719 (19.87%) | 24,967 (20.89%) |
| Never |  | 14,452 (48.84%) | 254,768 (46.83%) | 14,047 (48.81%) | 57,796 (48.36%) |
| Former |  | 9,082 (30.69%) | 174,291 (32.04%) | 8,843 (30.73%) | 36,754 (30.75%) |
| Unknown |  | 174 (0.59%) | 3,589 (0.66%) | - | - |
| Comorbidity |  |  |  |  |  |
| RA |  | 1,000 (3.38%) | 20,620 (3.79%) | 968 (3.39%) | 4,097 (3.43%) |
| OA |  | 3,125 (10.56%) | 61,163 (11.24%) | 3,032 (10.60%) | 12,647 (10.58%) |
| Anxiety |  | 10,276 (34.73%) | 180,583 (33.19%) | 9,922 (34.70%) | 40,929 (34.25%) |
| Depression |  | 12,786 (43.21%) | 233,067 (42.84%) | 12,329 (43.12%) | 51,161 (42.81%) |
| Schizophrenia |  | 604 (2.04%) | 12,681 (2.33%) | 579 (2.03%) | 2,559 (2.14%) |
| Alcohol dependence |  | 1,028 (3.47%) | 22,614 (4.16%) | 985 (3.45%) | 4,351 (3.64%) |
| Epilepsy |  | 842 (2.85%) | 16,771 (3.07%) | 810 (2.83%) | 3,430 (2.87%) |
| Substance Use Disorder |  | 873 (2.95%) | 20,662 (3.80%) | 835 (2.92%) | 3,756 (3.14%) |
| CCI |  |  |  |  |  |
| Low (0-2) |  | 20,535 (69.40%) | 367,848 (67.61%) | 19,753 (69.09%) | 82,688 (69.19%) |
| Medium (3-4) |  | 5,379 (18.18%) | 104,305 (19.17%) | 5,242 (18.33%) | 21,710 (18.16%) |
| High (≥5) |  | 3,675 (12.42%) | 71,897 (13.22%) | 3,597 (12.58%) | 15,119 (12.65%) |
| Concurrent drug use |  |  |  |  |  |
| Benzodiazepines |  | 6,915 (23.37%) | 123,889 (22.77%) | 6,662 (23.30%) | 27,439 (22.96%) |
| Antidepressants |  | 12,545 (42.40%) | 243,801 (44.81%) | 12,121 (42.39%) | 51,091 (42.75%) |
| Gabapentinoids |  | 120 (0.41%) | 2,980 (0.55%) | 117 (0.41%) | 543 (0.45%) |
| Muscle relaxants |  | 423 (1.43%) | 6,302 (1.16%) | 402 (1.41%) | 1,616 (1.35%) |
| Z-drugs |  | 3,403 (11.50%) | 65,917 (12.12%) | 3,290 (11.51%) | 13,747 (11.50%) |
| NSAIDs |  | 20,099 (67.96%) | 353,788 (65.03%) | 19,419 (67.92%) | 80,800 (67.61%) |

Note: L-TOT users= those who did not discontinue opioids; PS=propensity score; IMD = Index of Multiple Deprivation; MME= morphine milligram equivalent; RA: Rheumatoid Arthritis; OA: Osteoarthritis; CCI: Charlson Comorbidity Index; NSAIDs: Non-steroidal anti-inflammatory drugs

**Table 2.** Median years of follow-up and incidence rates of each outcome for L-TOT discontinuers and L-TOT users before matching.

| Outcome | Median years of follow-up (IQR) | Number of outcomes | Person-years | cIR per 10,000 person-years (95%CI) |
| --- | --- | --- | --- | --- |
| **Opioid-related death** |  |  |  |  |
| L-TOT discontinuers<br>(N=29,589) | 6.87 (3.32 to 11.51) | 28 | 220,278.56 | 1.27 (1.99 to 2.30) |
| Matched L-TOT users<br>(N=544,050) | 5.74 (2.51 to 10.18) | 740 | 3,457,487.2 | 2.14 (0.88 to 1.84) |
| **Hospitalisations due to bone fractures** |  |  |  |  |
| L-TOT discontinuers<br>(N=29,589) | 6.46 (3.01 to 11.09) | 2,791 | 211,391.22 | 132.03 (127.22 to 137.02) |
| Matched L-TOT users<br>(N=544,050) | 5.32 (2.16 to 9.76) | 50,755 | 3,371,142.5 | 150.56 (149.25 to 151.87) |
| **Composite outcome** |  |  |  |  |
| L-TOT discontinuers<br>(N=29,589) | 6.34 (2.94 to 10.99) | 2,812 | 209,244.15 | 134.39 (129.51 to 139.45) |
| Matched L-TOT users<br>(N=544,050) | 5.15 (2.02 to 9.57) | 51,424 | 3,298,540.1 | 155.90 (154.56 to 157.25) |
| **All-cause mortality** |  |  |  |  |
| L-TOT discontinuers<br>(N=29,589) | 7.30 (3.62 to 11.86) | 6,644 | 227,486.86 | 292.06 (285.12 to 299.17) |
| Matched L-TOT users<br>(N=544,050) | 6.02 (2.64 to 10.49) | 149,592 | 3,605,736.8 | 414.87 (412.78 to 416.98) |

Note: cIR=crude incidence rate; IQR=interquartile range; CI=confidence interval; Composite outcome=either opioid-related death or hospitalisations due to bone fractures or both

### Crude incidence rates

Over a median follow-up of 6.87 years (interquartile range [IQR]: 3.32 to 11.51) for L-TOT discontinuers and 5.74 years (IQR 2.51 to 10.18) for their matched L-TOT users, there were 28 opioid-related deaths in L-TOT discontinuers and 740 in L-TOT users, with a cIR of 1.27 (95% CI 1.99 to 2.30) and 2.14 (95% CI 0.88 to 1.84) per 10,000 person-years, respectively. In the analysis of hospitalisations due to bone fractures, which had a median follow-up of 6.46 years (IQR 3.01 to 11.09) in L-TOT discontinuers and 5.32 years (IQR 2.16 to 9.76) in L-TOT users, the incidence rate was lower among L-TOT discontinuers (132.03 per 10,000 person-years, 95% CI 127.22 to 137.02) compared to L-TOT users (150.56 per 10,000 person-years, 95% CI 149.25 to 151.87). A similar pattern was observed for the composite outcome, with cIRs of 134.39 (95% CI 129.51 to 139.45) per 10,000 person-years in L-TOT discontinuers and 155.90 (95% CI 154.56 to 157.25) per 10,000 person-years in L-TOT users, respectively. All-cause mortality was the most frequent adverse event, with 6,644 deaths in L-TOT discontinuers and 149,592 deaths in L-TOT users, corresponding to cIRs of 292.06 (95% CI 285.12 to 299.17) and 414.87 (95% CI 412.78 to 416.98) per 10,000 person-years, respectively, over a median follow-up of 7.30 years (IQR 3.62 to 11.86) in L-TOT discontinuers and 6.02 years (IQR 2.64 to 10.49) in L-TOT users.

### Adjusted hazards of each outcome

After adjusting for covariates used in the propensity score models, L-TOT discontinuation was associated with a 37% reduced risk in opioid-related death (aHR 0.63, 95% CI 0.42 to 0.94) (Table 3), and the PH assumption, tested after fitting the Cox model, was met for this outcome. However, for hospitalisations due to bone fractures, the composite outcome, and all-cause mortality, the assumption was met only during the initial follow-up period (approximately the first 0.3 years [ln(years) = -1]) but was violated at longer follow-up times (Supplementary Figure S5). To address this, time-dependent Cox proportional hazards models were applied, incorporating relevant covariates as time-dependent variables. The results showed that L-TOT discontinuation was associated with a 5% reduction in the risk of hospitalisations due to bone fractures (aHR 0.95, 95% CI 0.91 to 0.99), adjusting for age, epilepsy, CCI, NSAIDs, and antidepressants as time-dependent covariates. Similarly, L-TOT discontinuation was linked to a 7% reduced risk of the composite outcome (aHR 0.93, 95% CI 0.87 to 0.97), with age, CCI, and NSAIDs treated as time-dependent variables. For all-cause mortality, L-TOT discontinuation was associated with a 22% reduction of the risk (aHR 0.78, 95% CI 0.76 to 0.80), accounting for daily opioid dose (MME), osteoarthritis, anxiety, depression, NSAIDs, antidepressants, benzodiazepines, and Z-drugs as time-dependent factors (Table 3). Results from parametric survival models using a Weibull distribution were consistent with those from time-dependent Cox regression models (Table 3).

**Table 3.** Association between LTOT discontinuation and risks of each outcome in the 1:5 PSM cohorts.

| Cases and comparators | Adjusted HR (95%CI) based on Cox regression models | PH assumption test |  | Adjusted HR (95%CI) based on parametric survival analysis |
| --- | --- | --- | --- | --- |
|  |  | Covariate-specific test for L-TOT discontinuation | Global test for the full model |  |
| Opioid-related death |  |  |  |  |
| LTOT users (n=119,517) | 1.0 (ref) | P=0.4951 | P=0.4120 | N/A |
| LTOT discontinuers (n=28,592) | 0.63 (0.42 to 0.94) * |  |  |  |
| Hospitalisations due to bone fractures |  |  |  |  |
| LTOT users (n=120,236) | 1.0 (ref) | P=0.0774 | P<0.01 | 1.0 (ref) |
| LTOT discontinuers (n=28,650) | 0.95 (0.91 to 0.99) *.a |  |  | 0.96 (0.92 to 1.00) * |
| Composite outcome |  |  |  |  |
| LTOT users (n=120,389) | 1.0 (ref) | P=0.1074 | P<0.01 | 1.0 (ref) |
| LTOT discontinuers (n=28,654) | 0.93 (0.87 to 0.97) *.b |  |  | 0.96 (0.92 to 1.00) * |
| All-cause mortality |  |  |  |  |
| LTOT users (n=119,571) | 1.0 (ref) | P<0.01 | P<0.01 | 1.0 (ref) |
| LTOT discontinuers (n=28,503) | 0.78 (0.76 to 0.80) *.c |  |  | 0.80 (0.78 to 0.82) * |

Note: PSM=propensity score matched; CI=confidence interval; PH: proportional hazards; p>0.05 indicated that PH assumption was met; p<0.05 indicated that PH assumption was violated; \* =statistically significant; <sup>a</sup>=time-dependent cox regression adjusting for age, epilepsy, CCI, NSAIDs, and antidepressants as time-dependent covariates. <sup>b</sup>=time-dependent cox regression adjusting for age, CCI, and NSAIDs as time-dependent variables. <sup>c</sup> = time-dependent cox regression accounting for daily opioid dose (MME), osteoarthritis, anxiety, depression, NSAIDs, antidepressants, benzodiazepines, and Z-drugs as time-dependent variables.

Adjusted survival curves estimated from the Cox proportional hazards model for opioid-related death, shown in Figure 1, demonstrated a higher estimated survival probability in L-TOT discontinuers compared to L-TOT users throughout the follow-up period, consistent with the Cox regression result (aHR 0.65, 95% CI 0.62 to 0.68). However, it should be noted that, adjusted survival curves were not generated for the other three outcomes. This is because the hazard function in a time-dependent Cox model is influenced by covariates that change over time, making it infeasible to derive a single, unified survival curve.

**Figure 1.**
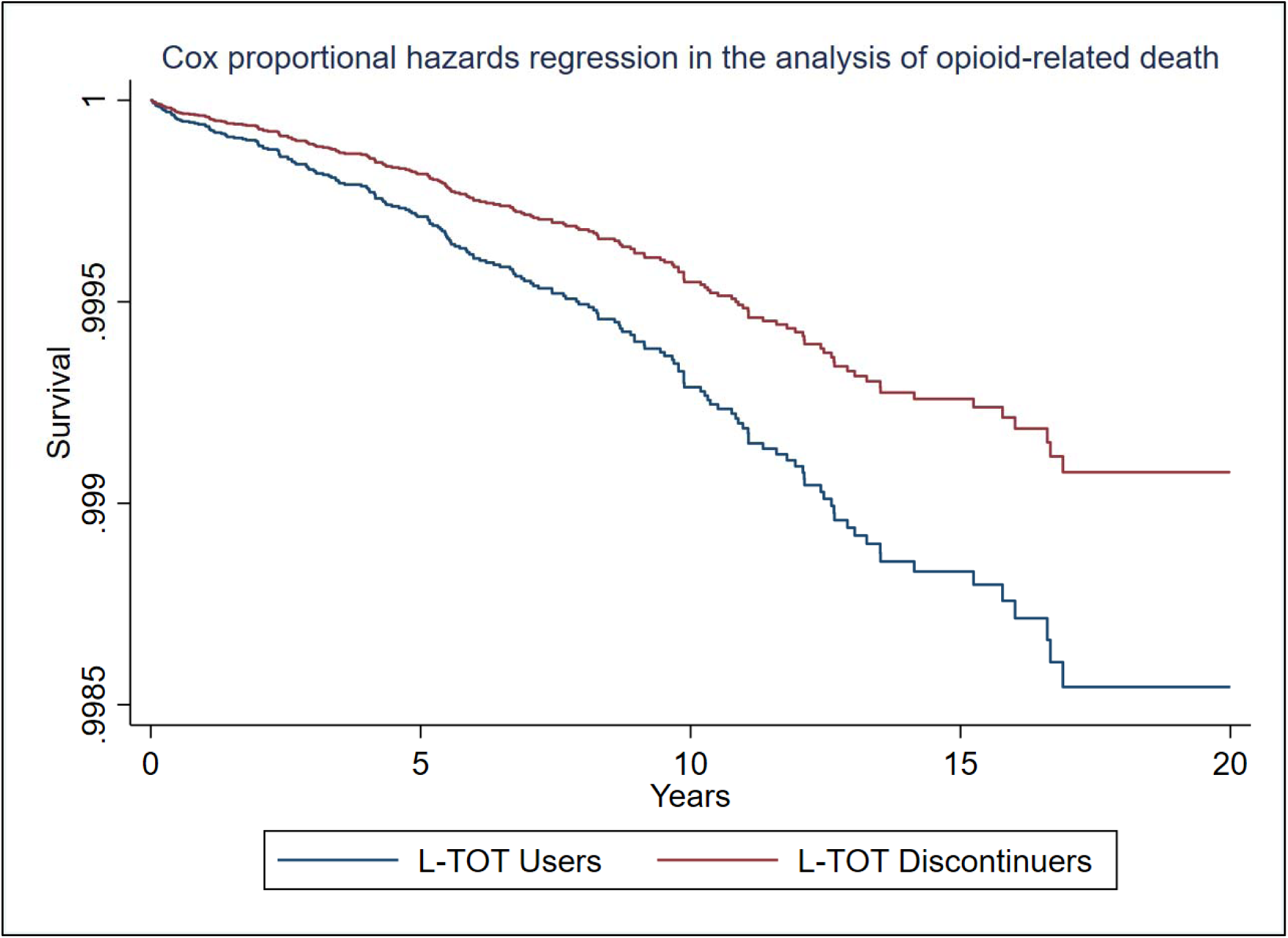
Adjusted survival curves based on Cox proportional hazards regression for opioid-related death between L-TOT discontinuers and L-TOT users in the 1:5 PSM cohort.

### Sensitivity analysis

In sensitivity analyses, regression results for each outcome in the 1:1 PSM cohorts were consistent with those in the 1:5 PSM cohorts, with aHRs (95% CI) of 0.65 (0.40 to 1.06) for opioid-related death, 0.94 (0.89 to 0.99) for hospitalisations due to bone fractures, 0.92 (0.89 to 0.97) for the composite, and 0.78 (0.75 to 0.80) for all-cause mortality, respectively (Supplementary Table S8). However, competing risk regressions, accounting for all-cause mortality as a competing event, found no significant associations between L-TOT discontinuation and opioid-related death (1:5 PSM: adjusted subhazard ratio [aSHR] 0.69, 95% CI 0.46 to 1.03; 1:1 PSM: aSHR 0.73, 95% CI 0.49 to 1.37), hospitalisations due to bone fractures (1:5 PSM: aSHR 1.05, 95% CI 1.01 to 1.10; 1:1 PSM: aSHR 1.05, 95% CI 1.00 to 1.11), or the composite outcome (1:5 PSM: aSHR 1.04, 95% CI 0.99 to 1.09; 1:1 PSM: aSHR 1.06, 95% CI 0.99 to 1.12) (Supplementary Table S9).

## Discussion

This population-based retrospective cohort study examined the association between L-TOT discontinuation and serious adverse outcomes among people with CNCP in UK primary care. Using PSM to balance covariates, we found that L-TOT discontinuation was significantly associated with reduced risks of opioid-related death, hospitalisations due to bone fractures, their composite and all-cause mortality. These findings were consistent across both 1:5 and 1:1 PSM analyses. However, when accounting for all-cause mortality as a competing risk, the associations between L-TOT discontinuation and the other outcomes (i.e., opioid-related death, hospitalisations due to bone fractures, and their composite) were attenuated and not statistically significant. This suggested that the observed safety benefits of discontinuation may be influenced by mortality from other causes.

### Comparison with other studies

Our study adds to the existing literature on L-TOT discontinuation outcomes by providing a unique perspective distinct from research primarily conducted in the US. Previous US-based studies have linked opioid discontinuation to increased mortality, particularly from overdose. For example, James et al. ^16^ found that patients who discontinued chronic opioid therapy had a nearly threefold higher risk of overdose death (aHR 2.94, 95% CI 1.01 to 8.61) compared to those who remained on opioids. Similarly, Fenton et al. ^30^ observed an increased risk of overdose (1.57, 1.42 to 1.74) up to two years after opioid taper (reduction). One possible explanation for these findings is that abrupt or forced discontinuation in the US may lead some individuals to seek opioids from alternative, potentially illicit sources, thus increasing the risk of overdose and suicide. Our study found that L-TOT discontinuation was associated with a reduced risk of opioid-related death and all-cause mortality. This discrepancy may stem from differences in patient characteristics, opioid prescribing practices, definitions of opioid discontinuation, and outcomes of interest. However, beyond these factors, opioid prescribing in the UK tends to follow a more conservative approach, characterised by lower initial daily doses, ^1^ and a closer clinical supervision, ^31^ which may reduce the likelihood of patients turning to illicit sources, and contribute to the lower incidence of overdose and mortality-related outcomes observed in our study.

Previous studies have reported increased psychological distress associated with the discontinuation or reduction of L-TOT. For example, Hallvik et al. ^13^ found a significantly higher risk of suicide following the discontinuation of chronic opioid therapy (aHR 3.63, 95% CI 1.42 to 9.25) compared to patients with stable or increasing doses. Larochelle et al. ^32^ observed an increased suicide risk with abrupt discontinuation (aHR 1.34, 95% CI 0.97 to 1.79) relative to stable dosage. Additionally, Fenton et al. ^30^ reported that opioid reduction was associated with increased rates of mental health crises (aHR 1.52, 95% CI 1.35 to 1.71) in two years after the initiation. Although our study did not directly assess suicide risk or mental health crises, the lower all-cause mortality and opioid-related death suggested that L-TOT discontinuation in UK primary care may not result in the same level of psychological distress reported in US studies. This difference may be attributed to the stronger integration of mental health services within the UK’s National Health Service (NHS), including programmes such as Improving Access to Psychological Therapies (IAPT) and the co-location of mental health practitioners within primary care settings. ^33, 34^ However, without direct measures of mental health outcomes, this remains a hypothesis that warrants further investigation.

Our study is one of the few that examined hospitalisations due to bone fractures following L-TOT discontinuation, finding a slight but significant reduction in the risk. While prolonged opioid use has been linked to increased fracture risk due to sedation, falls, and bone density loss, ^35^ previous studies have not consistently evaluated whether discontinuation reduces these risks. Our findings suggested that discontinuing L-TOT may confer modest benefits in this aspect for the CNCP population.

### Strengths and limitations

Our study has several strengths. First, to our knowledge, this is the first study in UK primary care examining serious adverse outcomes following L-TOT discontinuation in people with CNCP, using a large, nationally representative dataset (CPRD Aurum) with linked hospital and mortality records. These comprehensive datasets allowed for robust analyses of rare but clinically significant outcomes, such as opioid-related death, and enhanced the generalisability of our findings. Second, we employed rigorous PSM methods to improve the comparability between L-TOT discontinuers and L-TOT users, minimising potential confounding. The use of competing risk models to account for potential biases in mortality outcomes also ensured more accurate risk estimates. Finally, the median follow-up of over six years provided an opportunity to capture long-term effects of L-TOT discontinuation, providing insights beyond short-term withdrawal-related outcomes.

However, several limitations should be acknowledged. First, despite PSM, residual confounding may remain due to unmeasured factors, such as pain severity or psychosocial stressors, which could influence both discontinuation decisions and outcomes. Second, our study could not distinguish between patients who discontinued opioids due to clinical improvement versus those who discontinued due to adverse effects or provider-initiated discontinuation. This heterogeneity may have influenced outcome risks. Third, the observational design precludes causal inference, and we cannot rule out the possibility of reverse causality. Fourth, the reliance on electronic health records may have introduced misclassification bias, particularly for outcomes like bone fractures, which may not always result in hospitalisation. Fifth, L-TOT discontinuation was identified based on prescription records, which may not fully capture actual medication adherence or illicit opioid use. Sixth, the generalisability of our findings may be limited to similar healthcare systems, as prescribing practices and patient management strategies vary regionally. Finally, our study did not analyse the speed of discontinuation, nor did it focus on L-TOT reduction, which however, is more commonly practiced than discontinuation in clinic.

### Clinical implications

Current UK guidelines advocate for cautious opioid prescribing and regular reviews, recommending discontinuation when risks outweigh benefits. ^8, 36^ However, the lack of a standardised discontinuation (or reduction) approaches and patient support mechanisms makes the implementation of L-TOT discontinuation challenging, with concerns about withdrawal effects and patient resistance. ^31^ Despite this, our study provides evidence supporting L-TOT discontinuation, demonstrating its association with improved survival outcomes. This evidence suggests that primary care clinicians and prescribers could actively encourage L-TOT discontinuation when it is safe and feasible, particularly for patients who are concerned about pain management in the absence of opioids and may not initially foresee the benefits of discontinuation. ^37^ Recognising this, there is a clear need for targeted patient education programmes that emphasise the benefits of L-TOT discontinuation and actively involve patients in the process. For example, integrating shared decision-making models into clinical practice, such as recommended by NICE (National Institute for Health and Care Excellence), ^38^ can improve patient engagement and adherence to deprescribing protocols. However, it is important to note that, given the potential risks associated with abrupt discontinuation, e.g., suicide and overdose, ^13^ clinicians should adopt patient-centred, gradual reduction (or discontinuation) strategies with appropriate monitoring and supportive care.

Since our study found discontinuing L-TOT significantly reduces mortality risk, and evidence indicates that pain intensity does not worsen and may even slightly improve after discontinuation, ^10^ opioid discontinuation may be a safe and potentially beneficial option. However, it is crucial to identify specific patient groups for whom L-TOT discontinuation is most beneficial. When considering alternative pain management strategies, existing evidence supports the effectiveness of non-opioid interventions, such as CBT and physical exercise, ^39^ as they not only help manage pain but also enhance overall physical and mental well-being. Taken together, non-opioid treatments could serve as a viable and safer long-term approach to pain management, particularly for patients who may not require or benefit from prolonged opioid use. To support this transition, UK health policies should prioritise investment in multidisciplinary pain management services and improve access to non-opioid interventions, as advocated by NHS England’s Medicines Optimisation Strategy. ^40^

Finally, policymakers should incorporate updated real-world evidence into prescribing frameworks to ensure that clinical guidelines remain aligned with the latest research. The UK Government’s Overprescribing Review ^41^ acknowledges the need for evidence-driven policy changes, highlighting the importance of integrating emerging data such as ours into national opioid management strategies.

## Conclusions

Our study provides evidence that CNCP patients who discontinued L-TOT had a significant lower risk of opioid-related death, hospitalisations due to bone fractures, or all-cause mortality. These findings highlight the important safety benefits of L-TOT discontinuation. Future research should explore optimal opioid discontinuation strategies that balance harm reduction with adequate pain management. Additionally, studies focusing on other outcomes such as pain control, patient functional status, overall quality of life and healthcare utilisation following L-TOT discontinuation are needed to better understand the broader implications of opioid tapering and to guide evidence-based clinical decision-making.

## Ethical approval

This study was approved by the CPRD’s Independent Scientific Advisory Committee (protocol number: 23_002909, available at https://www.cprd.com/approved-studies/predictors-discontinuation-or-reduction-long-term-opioid-therapy-and-its). CPRD also has ethical approval from the Health Research Authority to support research using anonymised patient data (research ethics committee reference 21/EM/0265). Individual patient consent was not required as all data were deidentified. This study was reported according to the Strengthening the Reporting of Observational Studies in Epidemiology (STROBE) Statement (Supplementary Table S6). ^42^

## Data availability

This study used anonymised individual-level data from the CPRD Aurum. The data cannot be shared publicly due to CPRD regulations and ethical reasons. Other researchers can use the CPRD data in a secure environment by submitting a research protocol to the CPRD Independent Scientific Advisory Committee. Details of the application process and conditions of access are provided by the CPRD at https://www.cprd.com/Data-access.

## Acknowledgments

We appreciate Professor Daren Ashcroft and the NIHR Greater Manchester Patient Safety Research Collaboration, for covering the data access costs. We also thank all data providers and GPs for making anonymised data available for research.

## Contributors

QC conceived and designed the study. EK, TA, CG and YH supervised the conduct of the study. CM reviewed the code lists. QC conducted the data analysis and drafted the initial version of the manuscript. QC, EK, TA, CG, YH contributed to the interpretation of the findings. All authors have reviewed the manuscript and contributed to revisions. QC is the guarantor. The corresponding author (QC) attests that all listed authors meet authorship criteria and that no others meeting the criteria have been omitted.

## Transparency

The lead author (QC) confirms that the manuscript is an honest, accurate, and transparent account of the study, with no important omissions or unexplained discrepancies from the planned study.

## Dissemination to participants and related patient/public communities

As this study used anonymised CPRD data, direct dissemination to individuals is not possible. The University of Manchester networks will further disseminate the results, as the study forms a significant part of QC’s PhD.

## References

1. Cai Q, Huang Y-T, Allen T, Morris C, Grigoroglou C, Kontopantelis E. Trends of long-term opioid therapy and subsequent discontinuation among people with chronic non-cancer pain in UK primary care: a retrospective cohort study. medRxiv. 2025:2025.02.02.25321533.

2. Cai Q, Huang Y-T, Allen T, Morris C, Grigoroglou C, Kontopantelis E. Factors associated with long-term opioid therapy discontinuation for people with chronic non-cancer pain in UK primary care: a population-based retrospective cohort study. medRxiv. 2025:2025.03.19.25324292.

3. Burgstaller JM, Held U, Signorell A, Blozik E, Steurer J, Wertli MM. Increased risk of adverse events in non-cancer patients with chronic and high-dose opioid use—A health insurance claims analysis. PLoS One. 2020;15(9):e0238285.

4. Bedson J, Chen Y, Ashworth J, Hayward RA, Dunn KM, Jordan KP. Risk of adverse events in patients prescribed long-term opioids: a cohort study in the UK clinical practice research Datalink. European Journal of Pain. 2019;23(5):908–22.

5. Saunders KW, Dunn KM, Merrill JO, Sullivan M, Weisner C, Braden JB, et al. Relationship of opioid use and dosage levels to fractures in older chronic pain patients. Journal of general internal medicine. 2010;25:310–5.

6. Chen TC, Knaggs RD, Chen LC. Association between opioid-related deaths and persistent opioid prescribing in primary care in England: a nested case-control study. British Journal of Clinical Pharmacology. 2022;88(2):798–809.

7. Gomes T, Mamdani MM, Dhalla IA, Paterson JM, Juurlink DN. Opioid dose and drug-related mortality in patients with nonmalignant pain. Archives of internal medicine. 2011;171(7):686–91.

8. National Institute for Health and Care Excellence. NICE Guideline 193. Chronic pain (primary and secondary) in over 16s: assessment of all chronic pain and management of chronic primary pain (NG193). 2021. Available at: https://www.nice.org.uk/guidance/ng193.

9. Oxford University Hospitals, NHS foundation trust. Guidance for opioid reduction in primary care 2017. Available at: https://www.ouh.nhs.uk/services/referrals/pain/documents/gp-guidance-opioid-reduction.pdf.

10. McPherson S, Smith CL, Dobscha SK, Morasco BJ, Demidenko MI, Meath TH, et al. Changes in pain intensity after discontinuation of long-term opioid therapy for chronic noncancer pain. Pain. 2018;159(10):2097–104.

11. Frank JW, Lovejoy TI, Becker WC, Morasco BJ, Koenig CJ, Hoffecker L, et al. Patient outcomes in dose reduction or discontinuation of long-term opioid therapy: a systematic review. Annals of internal medicine. 2017;167(3):181–91.

12. Demidenko MI, Dobscha SK, Morasco BJ, Meath TH, Ilgen MA, Lovejoy TI. Suicidal ideation and suicidal self-directed violence following clinician-initiated prescription opioid discontinuation among long-term opioid users. General hospital psychiatry. 2017;47:29–35.

13. Hallvik SE, El Ibrahimi S, Johnston K, Geddes J, Leichtling G, Korthuis PT, et al. Patient outcomes after opioid dose reduction among patients with chronic opioid therapy. Pain. 2022;163(1):83–90.

14. Kennedy MC, Crabtree A, Nolan S, Mok WY, Cui Z, Chong M, et al. Discontinuation and tapering of prescribed opioids and risk of overdose among people on long-term opioid therapy for pain with and without opioid use disorder in British Columbia, Canada: A retrospective cohort study. Plos Medicine. 2022;19(12):e1004123.

15. Agnoli A, Xing G, Tancredi DJ, Magnan E, Jerant A, Fenton JJ. Association of dose tapering with overdose or mental health crisis among patients prescribed long-term opioids. Jama. 2021;326(5):411–9.

16. James JR, Scott JM, Klein JW, Jackson S, McKinney C, Novack M, et al. Mortality after discontinuation of primary care–based chronic opioid therapy for pain: a retrospective cohort study. Journal of general internal medicine. 2019;34(12):2749–55.

17. Gotthardt F, Huber C, Thierfelder C, Grize L, Kraenzlin M, Scheidegger C, et al. Bone mineral density and its determinants in men with opioid dependence. Journal of bone and mineral metabolism. 2017;35:99–107.

18. Clinical Practice Research Datalink (CPRD) Aurum March 2021 release notes. Available at: https://www.cprd.com/sites/default/files/2022-03/2022-03%20CPRD%20Aurum%20Release%20Notes.pdf

19. Jani M, Yimer BB, Sheppard T, Lunt M, Dixon WG. Time trends and prescribing patterns of opioid drugs in UK primary care patients with non-cancer pain: a retrospective cohort study. PLoS Medicine. 2020;17(10):e1003270.

20. Huang Y-T, Jenkins DA, Yimer BB, Jani M. Factors associated with long-term opioid use among patients with axial spondyloarthritis or psoriatic arthritis who initiate opioids. Rheumatology. 2024:keae444.

21. Morse S. Index of multiple deprivation (IMD)(UK). Encyclopedia of Quality of Life and Well-Being Research: Springer; 2024. p. 3440–3.

22. Lunt M, Pye S, Movahedi M. DrugPrep (STATA): an algorithm to transform RAW CPRD prescriptions data into formatted analysis-ready drug exposure data.(version v2. 0.0). Zenodo. 2018.

23. Nielsen S, Degenhardt L, Hoban B, Gisev N. A synthesis of oral morphine equivalents (OME) for opioid utilisation studies. Pharmacoepidemiology and drug safety. 2016;25(6):733–7.

24. Kanis JA, Johansson H, McCloskey EV, Liu E, Åkesson KE, Anderson FA, et al. Previous fracture and subsequent fracture risk: a meta-analysis to update FRAX. Osteoporosis international. 2023;34(12):2027–45.

25. Klotzbuecher CM, Ross PD, Landsman PB, Abbott III TA, Berger M. Patients with prior fractures have an increased risk of future fractures: a summary of the literature and statistical synthesis. Journal of bone and mineral research. 2000;15(4):721–39.

26. Herbert A, Wijlaars L, Zylbersztejn A, Cromwell D, Hardelid P. Data resource profile: hospital episode statistics admitted patient care (HES APC). International journal of epidemiology. 2017;46(4):1093-i.

27. Huang Y-q, Gou R, Diao Y-s, Yin Q-h, Fan W-x, Liang Y-p, et al. Charlson comorbidity index helps predict the risk of mortality for patients with type 2 diabetic nephropathy. Journal of Zhejiang University Science B. 2014;15:58–66.

28. Austin PC, Stuart EA. Moving towards best practice when using inverse probability of treatment weighting (IPTW) using the propensity score to estimate causal treatment effects in observational studies. Statistics in medicine. 2015;34(28):3661–79.

29. Chakrabarti A, Ghosh JK. AIC, BIC and recent advances in model selection. Philosophy of statistics. 2011:583–605.

30. Fenton JJ, Magnan E, Tseregounis IE, Xing G, Agnoli AL, Tancredi DJ. Long-term risk of overdose or mental health crisis after opioid dose tapering. JAMA network open. 2022;5(6):e2216726-e.

31. Public Health England. (2019). Dependence and withdrawal associated with some prescribed medicines: An evidence review. Available at: https://www.gov.uk/government/publications/prescribed-medicines-review-report.

32. Larochelle MR, Lodi S, Yan S, Clothier BA, Goldsmith ES, Bohnert AS. Comparative effectiveness of opioid tapering or abrupt discontinuation vs no dosage change for opioid overdose or suicide for patients receiving stable long-term opioid therapy. JAMA network open. 2022;5(8):e2226523-e.

33. National Health Service (NHS) Talking Therapies, for anxiety and depression. Available at: https://www.england.nhs.uk/mental-health/adults/nhs-talking-therapies/.

34. National Health Service (NHS) Mental Health Implementation Plan 2019/20 – 2023/24. Available at: https://www.longtermplan.nhs.uk/wp-content/uploads/2019/07/nhs-mental-health-implementation-plan-2019-20-2023-24.pdf.

35. Baldini A, Von Korff M, Lin EH. A review of potential adverse effects of long-term opioid therapy: a practitioner’s guide. The primary care companion for CNS disorders. 2012;14(3):27252.

36. Faculty of Pain Medicine. Opioids aware: a resource for patients and healthcare professionals to support prescribing of opioid medicines for pain. London: Faculty of Pain Medicine. Available at: http://www.fpm.ac.uk/faculty-of-pain-medicine/opioids-aware.

37. Nevedal AL, Timko C, Lor MC, Hoggatt KJ. Patient and provider perspectives on benefits and harms of continuing, tapering, and discontinuing long-term opioid therapy. Journal of General Internal Medicine. 2023;38(8):1802–11.

38. Shared decision making (NICE guideline NG197). Available at: https://www.sign.ac.uk/media/2008/20220927-nice-shared-decision-making.pdf.

39. Ehde DM, Dillworth TM, Turner JA. Cognitive-behavioral therapy for individuals with chronic pain: efficacy, innovations, and directions for research. American psychologist. 2014;69(2):153.

40. National Health Service (NHS) England. (2023). Medicines Optimisation Strategy: Improving the safe and effective use of medicines in the NHS. Available at: https://www.england.nhs.uk/medicines-optimisation.

41. Department of Health and Social Care. (2021). Good for you, good for us, good for everybody: A review of overprescribing in the NHS. Available at: https://assets.publishing.service.gov.uk/media/614a10fed3bf7f05ab786551/good-for-you-good-for-us-good-for-everybody.pdf.

42. Von Elm E, Altman DG, Egger M, Pocock SJ, Gøtzsche PC, Vandenbroucke JP. The Strengthening the Reporting of Observational Studies in Epidemiology (STROBE) statement: guidelines for reporting observational studies. The lancet. 2007;370(9596):1453-7.

